# Epidemiological Correlates of Overweight and Obesity in the Northern Cape Province, South Africa

**DOI:** 10.1101/2022.06.13.22276284

**Authors:** Mackenzie H Smith, Justin W Myrick, Oshiomah Oyageshio, Caitlin Uren, Jamie Saayman, Sihaam Boolay, Lena van der Westhuizen, Cedric Werely, Marlo Möller, Brenna M Henn, Austin W Reynolds

## Abstract

**Background:** In the past several decades, obesity has become a major public health issue worldwide, associated with increased rates of chronic disease and death. Like many developing nations, South Africa is experiencing rapid increases in BMI, and as a result, evidence-based preventive strategies are needed to reduce the increasing burden of overweight and obesity. This study aimed to determine the prevalence and predictors of overweight and obesity among a multi-ethnic cohort from the rural Northern Cape of South Africa.

**Methods:** These data were collected as part of a tuberculosis (TB) case-control study, with 395 healthy control participants included in the final analysis. Overweight and obesity were defined according to WHO classification. Multivariate linear models of BMI were generated using sex, age, education level, smoking, alcohol consumption, and diabetes as predictor variables. We also used multivariable logistic regression analysis to assess the relationship of these factors with overweight and obesity.

**Results:** The average BMI in our study cohort was 25.1. The prevalence overweight was 18.6% and the prevalence of obesity was 23.5%. We find that female gender, being older, and having more years of formal education are all positively associated with BMI in our dataset. Women (OR = 5.4, CI = 3.2-9.4), older individuals (OR = 1.02, CI = 1-1.04), and those with more years of education (OR = 1.18, CI = 1.07-1.29) were all more likely to be overweight or obese. Alternatively, being a smoker is negatively associated with BMI and decreases one’s odds of being overweight or obese (OR = 0.25, CI = 0.15-0.42).

**Conclusions:** We observed a high prevalence of overweight and obesity in this study. The odds of being overweight and obesity was higher in women and those with more education and increases with age. Community-based interventions to control obesity in these communities should pay special attention to these groups.

## Background

Obesity, a medical condition characterized by excessive fat accumulation, can have severe consequences for health, including increased risk of cardiovascular disease, cancer, stroke, and type 2 diabetes [1]. Obesity is most commonly determined through the measurement of Body Mass Index (BMI), a metric calculated by dividing an individual’s weight in kilograms by their height in meters squared. From 1990 to 2015, global mortality associated with elevated BMI increased by 28.3%, with the majority of these deaths being caused by cardiovascular disease [2]. In addition to its effects on individual health, high obesity incidence can have substantial economic implications, with a global impact of approximately $2.0 trillion annually [3]. The costs linked to obesity range from medical expenses and pharmaceuticals to absenteeism and premature mortality [4].

Global obesity prevalence increased almost threefold between 1975 and 2016 [5]. A 2015 study showed that 603.7 million adults globally were classified as obese, representing 12.0% of the adult population [2]. Based on current trends, it has been estimated that 1.12 billion adults worldwide will be classified as obese by 2030, and an additional 2.16 billion as overweight [6], roughly 40% of the world population. Sub-Saharan Africa in particular has shown staggering increases in obesity prevalence in the past several decades. Among seven countries in this region in 2009, an average of 31.4% of women were classified as overweight or obese [7]. In Malawi, 18% of men and 44% of women in urban areas were categorized as overweight or obese [8].

In accordance with both global trends and those observed in sub-Saharan Africa, the population of South Africa has also demonstrated a rapid increase in average BMI over the past several years [9–11]. As of 2002, 29.2% of men and 56.6% of women in South Africa were categorized as overweight or obese [12]. These values are higher than what has been reported in other African nations [13]. Various behavioral factors have also been studied in South African populations, with several studies indicating an inverse correlation between smoking and BMI [14, 15]. There is also evidence for higher obesity incidence among women as compared to men in South Africa, a trend often observed in developing nations [6, 13, 16].

It is important to note, however, that the majority of research on BMI and obesity in South Africa has been conducted on urban cohorts, particularly in the Eastern and Western Cape Provinces [17, 18]. Although national statistics on measures of obesity are available [19], research on rural and peri-urban South African areas, such as those in the Northern Cape Province, remains limited, restricting our understanding of how overweight and obesity manifest in these communities. This gap in the literature is important given the observed differences in obesity trends between urban and rural communities in lower- and middle-income nations [20]. In South Africa specifically, women in urban areas were 1.6 times more likely, and urban men 2.3 times more likely, to have excessive BMI than those in rural areas [16]. However, the average BMI of both men and women is increasing much more rapidly among rural communities than urban communities globally [21]. Further investigation in these smaller communities is crucial in identifying risk factors and working towards improved public health initiatives and education.

In this study, we analyze some of the demographic and behavioral factors related to BMI in the Northern Cape of South Africa. Our sample consists of 395 individuals from rural areas, small towns, and one large municipality in the province. Investigating these factors will lay the groundwork for the development of specialized public health and education initiatives to reduce the prevalence of obesity and overweight among this region.

## Methods

### Study Design and Sampling Procedure

The data used in this study was collected as part of the Northern Cape Tuberculosis (NCTB) Project, a case-control study on host susceptibility to TB. Between 2017 and the beginning of the SARS-CoV-2 pandemic in 2020, data was collected on 1,095 individuals (N_men_ = 544; N_women_ = 551) recruited from 12 community (public) health clinics in the Northern Cape Province, South Africa that serve populations with high TB rates, mostly in rural towns with populations under 10,000. The Northern Cape Province has the largest area, lowest population size and lowest population density in South Africa. Data was collected via participant interviews, medical histories, saliva samples, and anthropometric measurements. In this study, only participants with a negative TB result were included due to a symptomatic effect of TB on BMI [22]. Participants were partitioned into the case or control group based on a decision tree considering previous TB diagnosis, treatment for TB, and TB test results obtained during the course of the study (Oyageshio, in prep.).

### Ethics and Informed Consent

Data collected from study participants was approved by the Health Research Ethics Committee of Stellenbosch University (Project number: N11/07/210A) and the Institutional Review Board of the University of California, Davis (IRB number: 1749073-1). Participation in this study was voluntary, with the ability to withdraw at any time. Written informed consent was obtained and subsequent medical and demographic questionnaires were conducted in the local language of Afrikaans by trained research assistants from the community. All data was kept confidential with no connections to participant names. Deidentified variables were stored in a secure RedCap database following data collection.

### Demographic and Socio-economic Factors

Demographic information was collected through interviews and recorded on a data collection sheet. Participants were asked to provide their town of residence, highest level of education achieved, and the ethnic group with which they self-identify.

### Medical History

During interviews, participants self-reported diabetes, HIV, asthma, and TB status. If participants reported having diabetes, they were asked to identify if they were a type 1 or type 2 diabetic. HIV status was self-reported as positive, negative, or unknown. Asthma was recorded as no, yes, or unknown. In addition to asking participants whether they had TB at the time of the study, they were also asked if they were currently taking TB medication, whether they had TB in the past, and if so, how many TB episodes they had experienced. Following the self-reporting of TB information, TB tests were administered to all participants using GeneXpert, Auramine O Stain, GeneXpert Ultra, SMEAR, or Culture tests.

### Behavioral Factors

Alcohol use was evaluated by asking participants if they drink alcohol. If yes, further information was collected regarding the specifics of their alcohol consumption. Participants were asked whether they drink beer, wine, liquor, or ginger beer, and how much of each they consume during the week and weekend. Smoking behavior was categorized as yes or no. If participants answered yes to smoking, they were asked to provide the age at which they began smoking, as well as the average amount they smoke per day.

### Anthropometry

Height was measured using a stadiometer. Participants were asked to look straight ahead and stand with their heels against the wall while measurements were being taken. One height measurement was recorded for participants in pilot data collection and two measurements were taken upon initiation of the primary study. For individuals with two recorded measurements for height, the average of these values was used. Weight was measured with a digital scale and recorded once. Height and weight measurements were used to calculate BMI for each participant using the equation: BMI = kg/m^2^. Waist and hip circumference measurements were self-administered by participants using a measuring tape, and two measurements of each were recorded. Following calculation of BMI, participants were assigned to weight categories based on the guidelines set forth by the Centers for Disease Control and Prevention: those with a BMI < 18.5 were classified as “Underweight”, 18.5 to <25 as “Healthy,” 25 to <30 as “Overweight,” and 30 or higher as “Obese” [23]. Waist-to-hip ratio was calculated by dividing average waist circumference (cm) by average hip circumference (cm).

### Quality Control

Participants who were TB positive, HIV positive, or both (N = 642), were removed from this study due to an expected effect of disease on BMI. All hip and waist measurements supervised by community health care worker #5 were also removed due to errors in measurement technique. BMI could not be calculated for an additional 18 participants due to an absence of height measurements, weight measurements, or both, and these individuals were removed. A total of 395 individuals were included in BMI analyses.

### Statistical Analyses

Quality control and data analysis were performed in R 4.0.2. The packages effects [24, 25] was used for data analysis, and ggplot2 [26] was used for data visualization.

Six covariates (alcohol intake (yes or no), smoking (yes or no), diabetes (yes or no), sex, age, and years of education) were entered into a generalized linear model to further characterize their association with BMI. As observed in other studies, BMI was not normally distributed among our sample, therefore we used log-transformed BMI as the outcome variable. Bivariate effect plots were generated between BMI and each individual covariate included in the model. Additional generalized linear models were generated by partitioning male and female participants to examine sex-specific effects.

We also performed statistical analyses using waist-to-hip ratio (WHR) in place of BMI. A generalized linear model was created using WHR as the outcome variable and included alcohol intake, smoking, diabetes, sex, years of education, and age. This model included 161 participants. Bivariate effect plots were created between WHR and alcohol intake, smoking, diabetes, sex, age, and highest qualification. Our generalized linear models were supplemented by the generation of odds ratios using the questionr package [27] to investigate the relationship between individual variables and BMI outcome (overweight/obese versus not).

## Results

### Participant Demographics

Our final sample (**Table 1**) included 395 participants (n_male_ = 145; n_female_ = 250). The median age was 43 years, with all participants falling between 18 and 86 years of age. Our dataset was collected across twelve study sites, representing rural areas, small towns, and one small city from the Northern Cape Province, South Africa. The most common self-identified ethnicity among participants was Coloured (85%), followed by Tswana (5%). Approximately 7% of participants reported having no education, 27% reported attending or completing primary school, and 66% reported attending or completing secondary school.

**Table 1:** Summary of participant demographics, behavioral factors, anthropometry, and BMI. Summary statistics are provided for men, women, and the total sample. We find substantial differences in average BMI and overweight/obesity incidence between men and women.

| Variable | Men | Women | Total |
| --- | --- | --- | --- |
| Participants (N) | 145 | 250 | 395 |
| Age (mean) | 44 | 43 | 43 |
| Years of education (mean) | 8 | 8 | 8 |
| Smokes | 75% | 54% | 62% |
| Drinks Alcohol | 50% | 34% | 39% |
| Height (cm) (mean) | 165.2 | 155.7 | 159.2 |
| Weight (kg) (mean) | 57 | 67.3 | 63.5 |
| BMI (mean) | 21 | 27.8 | 25.3 |
| WHR (mean)* | 0.84 | 0.88 | 0.87 |
| Underweight | 39% (n=57) | 12% (n=31) | 22% (n=88) |
| Healthy weight | 43% (n=62) | 31% (n=77) | 35% (n=139) |
| Overweight | 13% (n=19) | 21% (n=52) | 18% (n=71) |
| Obese | 5% (n=7) | 36% (n=90) | 25% (n=97) |

### Behavioral Factors

The majority (62%) of participants were smokers. Among women, 54% reported that they smoked, compared to 75% of men. Only 39% of participants reported consuming some quantity of alcohol. This also differed by sex, with 50% of men and 34% of women in our sample reporting alcohol consumption.

### Anthropometry and BMI

Median height and weight were 159cm (range: 109.4-184.9cm) and 60kg (range: 27-134kg) respectively. The median BMI was 23.6, and the mean BMI was 25.3. Nearly a quarter (22%) of participants were classified as underweight, 35% as healthy, 18% as overweight, and 25% as obese. The distribution of individuals between these classifications differed substantially between male and female participants, with 36% of female participants being placed in the obese category compared to only 5% of male participants (**Figure 1; Table 1**).

**Figure 1:**
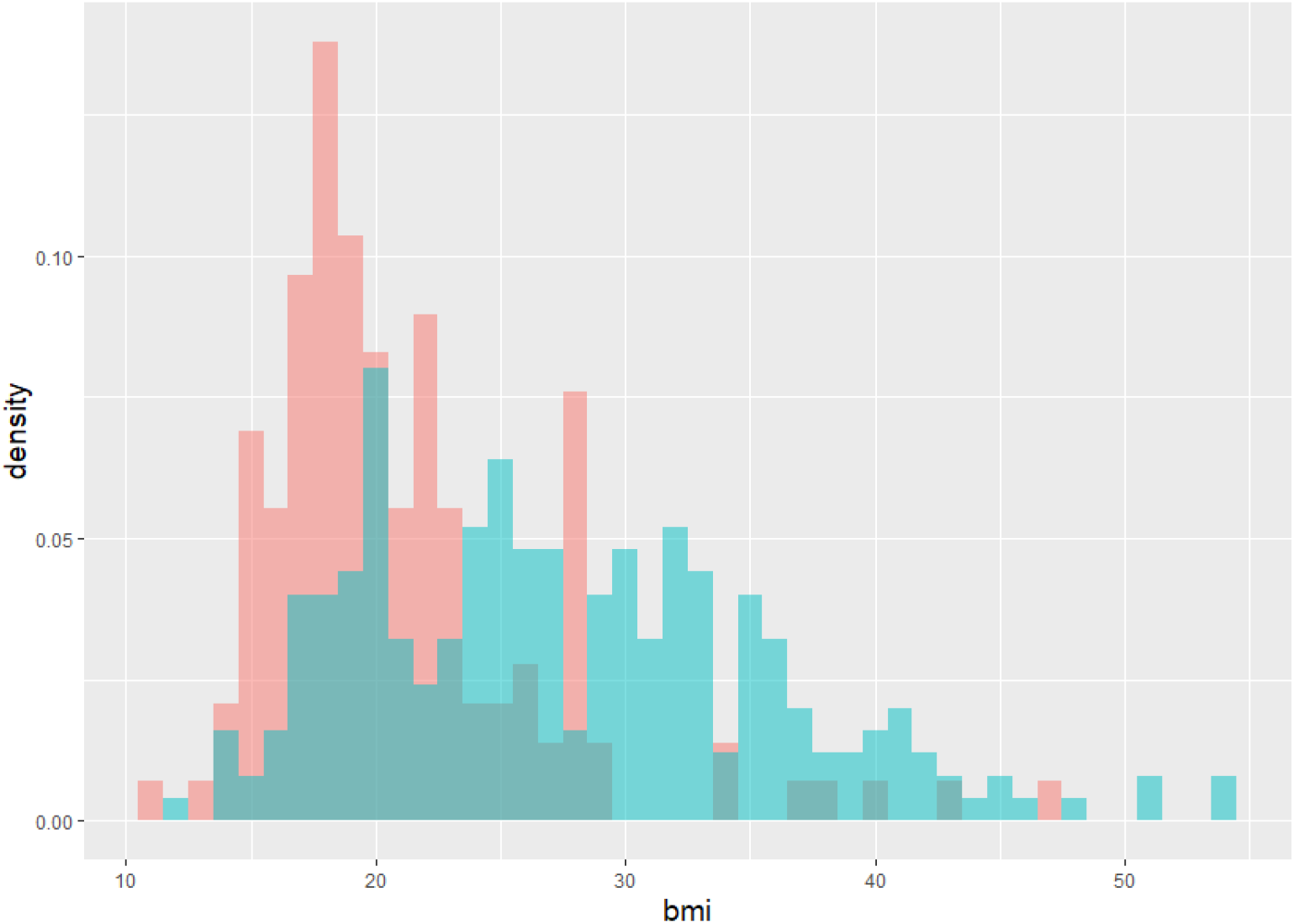
Histogram of BMI values in our dataset, partitioned by sex. Pink bars represent the distribution in males, while blue bars represent the distribution in females. We find that the average BMI for females is higher than that of males.

### Factors associated with BMI, WHR, and obesity

We fit a generalized linear model to investigate the relationship between BMI and the factors of sex, age, education, smoking, drinking, and diabetes status. This model showed significant relationships between BMI and smoking, gender, education level, age, and diabetes (**Table 2**). Together, these factors explained 26% of the variance in BMI among our cohort. Being female showed a strong positive correlation with BMI, as did age and years of education. The relationships between each of these variables and BMI were observed in our bivariate effect plots (**Figure 2**), which indicated a 5-point BMI increase for women, a 4-point decrease for smoking, and a 6-point increase for having 14 years of education as compared to zero.

**Table 2:** Model 1 results showing the impact of six covariates on BMI outcomes in our sample of 395 participants. We find significant positive correlations between BMI and age, years of education, and being female. We find a significant negative correlation between smoking and BMI.

| Variable | Estimate | se | z | Pr(>\|z\|) |
| --- | --- | --- | --- | --- |
| (Intercept) | -4.866689 | 0.943656 | -5.157 | 2.51E-07 |
| Alcohol | 0.214465 | 0.268316 | 0.799 | 0.42412 |
| Sex | 1.687443 | 0.275395 | 6.127 | 8.94E-10 |
| Age | 0.023191 | 0.009507 | 2.439 | 0.01471 |
| Years of education | 0.162407 | 0.047261 | 3.436 | 0.00059 |
| Smokes | -1.372207 | 0.256907 | -5.341 | 9.23E-08 |
| Diabetes | 1.440148 | 0.577864 | 2.492 | 0.0127 |

**Figure 2:**
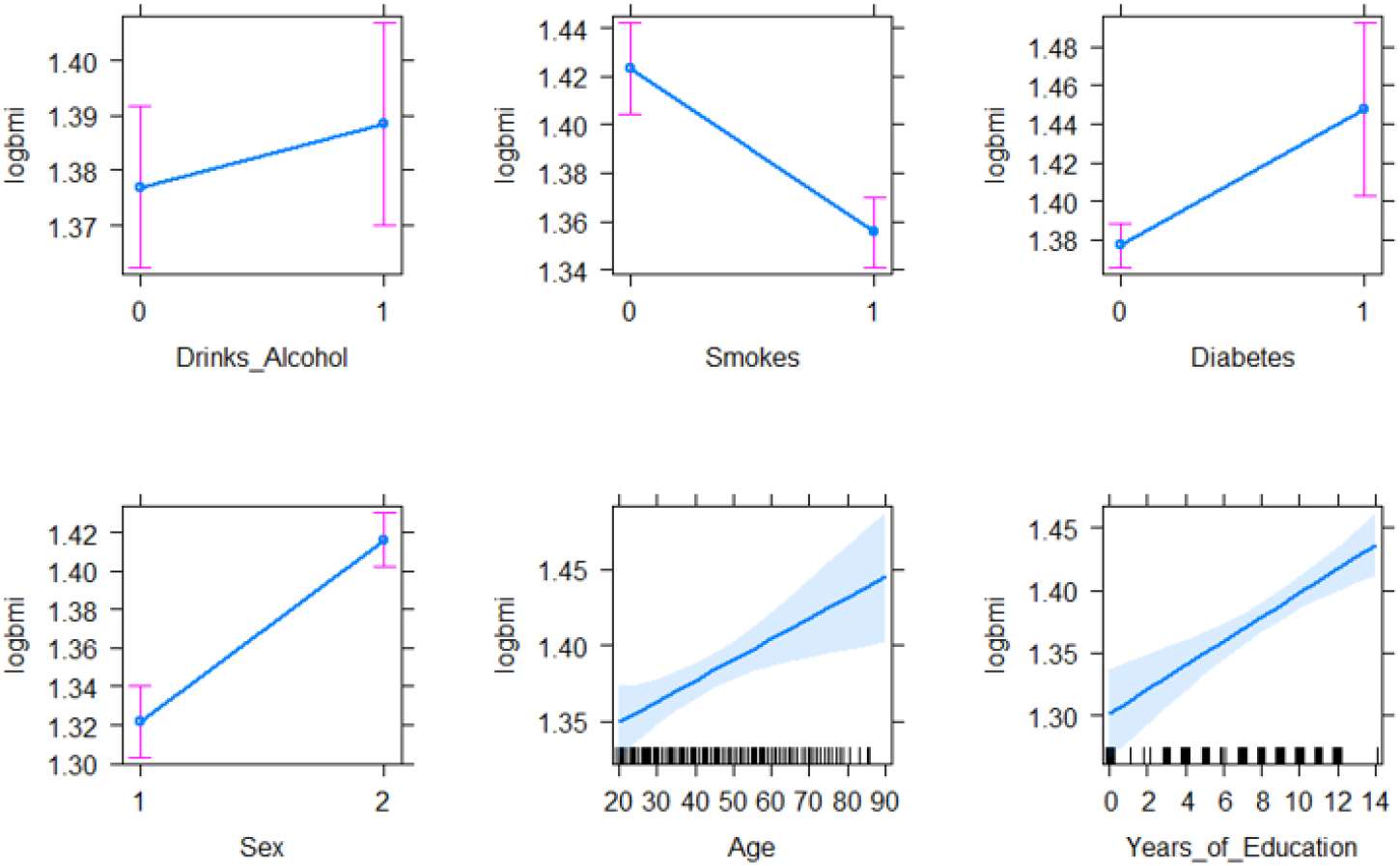
Bivariate effect plots demonstrating the relationship between BMI and the covariates included in Model 1. We find a 5-point BMI increase associated with being female, a 6-point increase for having 14 years of education, and a 4-point decrease for smoking.

We find evidence that women in our sample had over 5 times greater odds of being overweight or obese compared to men (OR = 5.4, CI = 3.2-9.4; **Table 3**). In our dataset, every one-year increase in age is associated with a 2% increase in the odds of being overweight or obese, while each additional year of school completed increased those odds by 18%. Smoking, on the other hand, showed a significant negative correlation with BMI (p = 5.85×10^−8^). In fact, smoking decreased the odds of being overweight or obese by nearly 75% in our study population (OR = 0.25, CI = 0.15-0.42).

**Table 3:** Odds ratio results for the covariates included in Model 1. We find that women have 5x greater risk of being overweight or obese, each additional year of school increases risk by 18%, and smoking decreases risk by 75%.

| Variable | OR | Lower CI (2.5%) | Upper CI (97.5%) | p |
| --- | --- | --- | --- | --- |
| (Intercept) | 0.0076988 | 0.0011427 | 0.0466 | 2.51E-07 |
| Alcohol | 1.2391987 | 0.7352237 | 2.1097 | 0.4241174 |
| Sex | 5.4056422 | 3.1941477 | 9.4323 | 8.94E-10 |
| Age | 1.0234617 | 1.0047186 | 1.043 | 0.0147148 |
| Years of education | 1.1763391 | 1.074636 | 1.294 | 0.0005895 |
| Smokes | 0.2535469 | 0.1520819 | 0.4172 | 9.23E-08 |
| Diabetes | 4.221319 | 1.4584546 | 14.5498 | 0.0126958 |

Because of the strong sex effect on BMI, we also fit models for women (Table S1; n = 250) and men (Table S2; n = 145) separately to investigate potential sex-specific factors associated with BMI. The strong negative correlation between BMI and smoking seen in our primary model was maintained in both sex-specific models (p = 4.4×10-5 and 3.2×10-3, respectively). The significant positive correlations between years of education, age, diabetes and BMI are maintained with the women-only model, however these relationships were notably weaker than in the primary model. None of these significant correlations are seen in the men-only model.

We also explored the factors associated with waist-to-hip measurement ratios in our dataset. This model was fit on a subset of the dataset (Table S3; n=160), as waist and hip measurements were not available for all participants. We find a slight positive correlation between WHR and years of education (p = 0.0412), but together these variables only explain ∼2% of the variance in WHR in our dataset. No other covariates showed a significant association with WHR.

## Discussion

In this study we analyzed the demographic and socio-behavioral factors related to BMI in an adult cohort from the Northern Cape Province, South Africa. Our results suggest a significant difference in obesity trends between men and women in the study population. Sex-based differences were observed not only in obesity incidence, but also in the degree to which various factors impact BMI outcomes. While smoking has a strong negative correlation with BMI in both men and women, we find that age and years of education are only significantly correlated with BMI in women. Although we had significantly more female participants (250 women, 145 men), our results are consistent with previous research. Higher obesity incidence among women as compared to men is observed in other studies on South African populations [9, 14–16]. While data regarding the effect of socioeconomic status on BMI outcomes in males and females shows variability depending on the metric used to represent socioeconomic status, Wagner et al. found that primary and tertiary education were associated with greater BMI values in females only [14]. Interestingly, while we find an association between age and BMI among females only, previous research has demonstrated this correlation for males only [15, 16].

One limitation of the current study is the relatively limited sample size of waist and hip measurements. While BMI is a widely used metric for obesity that is correlated with body fat content, past research has shown that researchers must be careful not to over-interpret BMI in studies of obesity, as it does not distinguish between excess fat, muscle, or bone mass, or provide any indication of the distribution of fat among individuals [28, 29]. Several studies have suggested that WHR are better at predicting visceral adiposity and cardiometabolic disease risk [30, 31]. Future work incorporating these and other more sensitive measures of adiposity will provide greater detail into the risk-factors and consequences of obesity in this population.

In line with prior research on sex differences in BMI and obesity, we find 57% of female participants were overweight or obese compared to only 19% of male participants. One such study found that in 5 out of 6 sampling locations in sub-Saharan Africa, a higher percentage of females were categorized as obese as compared to males [32]. A study in Soweto, South Africa showed that the proportion of women classified as obese was twice that of men [15]. There are several potential explanations for the extreme obesity risk associated with being female in our sample population. There is some evidence to suggest higher BMI is desirable in some South African communities [33, 34]. It has also been shown that having control over household food spending is associated with greater incidence of obesity in women [33].

Previous work has shown that socioeconomic status, including educational level, is positively correlated with BMI, for both men and women, in South Africa [32, 35, 36], as well as other low- and middle-income nations [32, 37]. This aligns with our result demonstrating the more education one has, the higher their BMI – each year of education increases their odds of being overweight or obese by 18%. This trend may be explained by cultural factors, as being overweight or obese reflects wealth in low-income communities where many lack adequate access to food. A study on 37 lower- and middle-income nations showed that BMI was positively associated with wealth in all 37 countries surveyed [38]. Previous work has also found that a person’s daily amount of vigorous physical activity also decreases with increasing wealth [39, 40], suggesting a multifactorial relationship between socioeconomic status and BMI. Interestingly, we do not see a significant relationship between age and years of education in our model of men-only. This pattern has been reported elsewhere [33] and suggest that other factors may be more important in explaining BMI variation in men in the Northern Cape.

Our results indicating that smoking decreases odds of being overweight or obese by 75% are also in line with previous research. Nicotine use suppresses appetite and causes an increased resting metabolic rate, resulting in weight loss [41]. Smoking has previously been associated with lower BMI in South African adults [15, 32].

## Conclusion

Overall, our results suggest that smoking, sex, education level, age, and diabetes all influence BMI outcomes among the rural population of the Northern Cape of South Africa, consistent with previous research in other South African cohorts and developing nations. These results deepen our understanding of the factors contributing to obesity risk in this region. A necessary future direction of this work will be to include additional measures of adiposity, behavior, and socio-economic status to obtain a more complete understanding of the risk-factors for obesity in the Northern Cape. This will allow for better identification of high-risk groups for behavioral interventions, ultimately mitigating the public health and economic burdens created by the global obesity epidemic at the local level.

## Data Availability

The data that support the findings of this study are available from the authors but restrictions apply to the availability of these data, due to the constraints of the informed consent, and so are not publicly available. Data are however available from the authors upon reasonable request and with permission of the Human Research Ethics Committee at Stellenbosch University.

## Declarations

### Ethics approval and consent to participate

Data was collected from participants in accordance with the Declaration of Helsinki and was approved by the Health Research Ethics Committee of Stellenbosch University (Project number: N11/07/210A) and the Institutional Review Board of the University of California, Davis (IRB number: 1749073-1). Written informed consent was obtained from all participants.

### Consent for publication

Not applicable

### Author Contributions

MM, BMH, CW, and AWR conceived the study. JWM, LW, JS, SB, BMH, and AWR contributed to data collection. MHS, JWM, JS, SB, CU, OO, and AWR contributed to data processing and analysis. MHS and AWR wrote the manuscript with editing from all other authors. All authors read and approved the final manuscript.

## Acknowledgements

First and foremost, we would like to thank our participant communities in the Northern Cape for their continued trust and support in helping us undertake this project. We would also like to thank our community research assistants and translators who assisted in data collection for the project. Finally, we want to thank the Department of Health in the Northern Cape Province, South Africa for their continued support of the project.

## References

1. CDC. Adult Obesity Facts. 2021.

2. The GBD 2015 Obesity Collaborators. Health Effects of Overweight and Obesity in 195 Countries over 25 Years. N Engl J Med. 2017;377:13–27.

3. Dobbs R, Sawers C, Thompson F, Manyika J, Woetzel J, Child P, et al. Overcoming obesity: An initial economic analysis. 2014.

4. Dee A, Kearns K, O’Neill C, Sharp L, Staines A, O’Dwyer V, et al. The direct and indirect costs of both overweight and obesity: a systematic review. BMC Res Notes. 2014;7:242.

5. WHO. Obesity and Overweight Factsheet. 2021.

6. Popkin BM, Adair LS, Ng SW. Global nutrition transition and the pandemic of obesity in developing countries. Nutrition Reviews. 2012;70:3–21.

7. Ziraba AK, Fotso JC, Ochako R. Overweight and obesity in urban Africa: A problem of the rich or the poor? BMC Public Health. 2009;9:465.

8. Price AJ, Crampin AC, Amberbir A, Kayuni-Chihana N, Musicha C, Tafatatha T, et al. Prevalence of obesity, hypertension, and diabetes, and cascade of care in sub-Saharan Africa: a cross-sectional, population-based study in rural and urban Malawi. The Lancet Diabetes & Endocrinology. 2018;6:208–22.

9. Cois A, Day C. Obesity trends and risk factors in the South African adult population. BMC Obes. 2015;2:42.

10. Steyn K, Fourie J, Rossouw JE, Langenhoven ML, Joubert G, Chalton DO. Anthropometric profile of the coloured population of the Cape Peninsula. 1990;78:5.

11. Steyn K, Bourne L, Jooste P, Fourie J, Rossouw K, Lombard C. Anthropometric profile of a black population of the Cape Peninsula in South Africa. East African Medical Journal. 1998;75:35–40.

12. Puoane T, Steyn K, Bradshaw D, Laubscher R, Fourie J, Lambert V, et al. Obesity in South Africa: The South African Demographic and Health Survey. Obesity Research. 2002;10:1038– 48.

13. Goedecke J, Jennings C, Lambert E. Obesity in South Africa. Chronic Diseases of Lifestyle in South Africa: 1995-2005. 2006;:65–79.

14. Wagner RG, Crowther NJ, Gómez-Olivé FX, Kabudula C, Kahn K, Mhembere M, et al. Sociodemographic, socioeconomic, clinical and behavioural predictors of body mass index vary by sex in rural South African adults-findings from the AWI-Gen study. Global Health Action. 2018;11:1549436.

15. Micklesfield LK, Kagura J, Munthali R, Crowther NJ, Jaff N, Gradidge P, et al. Demographic, socio-economic and behavioural correlates of BMI in middle-aged black men and women from urban Johannesburg, South Africa. Global Health Action. 2018;11:1448250.

16. Okop KJ, Levitt N, Puoane T. Factors Associated with Excessive Body Fat in Men and Women: Cross-Sectional Data from Black South Africans Living in a Rural Community and an Urban Township. PLoS ONE. 2015;10:e0140153.

17. Malhotra R, Hoyo C, Østbye T, Hughes G, Schwartz D, Tsolekile L, et al. Determinants of obesity in an urban township of South Africa. South African Journal of Clinical Nutrition. 2008;21:315–20.

18. Owolabi EO, Ter Goon D, Adeniyi OV. Central obesity and normal-weight central obesity among adults attending healthcare facilities in Buffalo City Metropolitan Municipality, South Africa: a cross-sectional study. J Health Popul Nutr. 2017;36:54.

19. Statistics South Africa, South Africa, editors. South Africa Demographic and Health Survey 2016. Key indicators report. Pretoria: Statistics South Africa; 2017.

20. Neuman M, Kawachi I, Gortmaker S, Subramanian SV. Urban-rural differences in BMI in low-and middle-income countries: the role of socioeconomic status. The American Journal of Clinical Nutrition. 2013;97:428–36.

21. NCD Risk Factor Collaboration (NCD-RisC). Rising rural body-mass index is the main driver of the global obesity epidemic in adults. Nature. 2019;569:260–4.

22. Zhang H, Li X, Xin H, Li H, Li M, Lu W, et al. Association of Body Mass Index with the Tuberculosis Infection: a Population-based Study among 17796 Adults in Rural China. Sci Rep. 2017;7:41933.

23. CDC.Defining Adult Overweight and Obesity. 2021.

24. Fox J. E□ect Displays in R for Generalised Linear Models.pdf. Journal of Statistical Software. 2003;8.

25. Fox J, Weisberg S. An R Companion to Applied Regression. 3rd Edition. Thousand Oaks, CA: Sage; 2019.

26. Wickham H. ggplot2: Elegant Graphics for Data Analysis. New York, NY: Springer; 2016.

27. Barnier J, Briatte F, Larmarange J. questionr: Functions to Make Surveys Processing Easier. 2022.

28. Rothman KJ. BMI-related errors in the measurement of obesity. Int J Obes. 2008;32:S56–9.

29. Adab P, Pallan M, Whincup PH. Is BMI the best measure of obesity? BMJ. 2018;:k1274.

30. Song X, Jousilahti P, Stehouwer CDA, Söderberg S, Onat A, Laatikainen T, et al. Comparison of various surrogate obesity indicators as predictors of cardiovascular mortality in four European populations. Eur J Clin Nutr. 2013;67:1298–302.

31. Lee CMY, Huxley RR, Wildman RP, Woodward M. Indices of abdominal obesity are better discriminators of cardiovascular risk factors than BMI: a meta-analysis. Journal of Clinical Epidemiology. 2008;61:646–53.

32. Ramsay M, Crowther NJ, Agongo G, Ali SA, Asiki G, Boua RP, et al. Regional and sex-specific variation in BMI distribution in four sub-Saharan African countries: The H3Africa AWI-Gen study. Global Health Action. 2018;11:1556561.

33. Case A, Menendez A. Sex differences in obesity rates in poor countries: Evidence from South Africa. Economics & Human Biology. 2009;7:271–82.

34. Okop KJ, Mukumbang FC, Mathole T, Levitt N, Puoane T. Perceptions of body size, obesity threat and the willingness to lose weight among black South African adults: a qualitative study. BMC Public Health. 2016;16:365.

35. Alaba O, Chola L. Socioeconomic Inequalities in Adult Obesity Prevalence in South Africa: A Decomposition Analysis. IJERPH. 2014;11:3387–406.

36. Wandai ME, Aagaard-Hansen J, Manda SO, Norris SA. Transitions between body mass index categories, South Africa. Bull World Health Organ. 2020;98:878–885I.

37. Dinsa GD, Goryakin Y, Fumagalli E, Suhrcke M. Obesity and socioeconomic status in developing countries: a systematic review. Obesity Reviews. 2012;13:1067–79.

38. Neuman M, Finlay JE, Davey Smith G, Subramanian S. The poor stay thinner: stable socioeconomic gradients in BMI among women in lower-and middle-income countries. The American Journal of Clinical Nutrition. 2011;94:1348–57.

39. Gradidge PJ-L, Norris SA, Munthali R, Crowther NJ. Influence of socioeconomic status on changes in body size and physical activity in ageing black South African women. Eur Rev Aging Phys Act. 2018;15:6.

40. Micklesfield LK, Pedro TM, Kahn K, Kinsman J, Pettifor JM, Tollman S, et al. Physical activity and sedentary behavior among adolescents in rural South Africa: levels, patterns and correlates. BMC Public Health. 2014;14:40.

41. Audrain-McGovern J, Benowitz NL. Cigarette Smoking, Nicotine, and Body Weight. Clin Pharmacol Ther. 2011;90:164–8.

